# Gray matter morphology and pain-related disability in young adults with low back pain

**DOI:** 10.1101/2024.05.29.24308150

**Authors:** Jo Armour Smith, Rongwen Tain, Isaac Chrisman, Kelli G. Sharp, Laura M Glynn, Linda R. Van Dillen, Jesse V. Jacobs, Steven C. Cramer

## Abstract

Structural neuroplasticity in the brain may contribute to the persistence of low back pain (LBP) symptoms and the disability associated with them. It is not known if structural adaptations are evident early in the lifespan in young adults with LBP. This study compared gray matter in cortical sensorimotor regions in young adults with and without persistent LBP and identified gray matter and clinical predictors of pain-related disability. Eighty-two individuals with and without a history of LBP participated. Peak and average gray matter density in cortical sensorimotor regions of interest was quantified using voxel-based morphometry. Pain-related disability, pain intensity, pain duration, and pain-related fear were also assessed. Multiple linear regression was used to determine independent predictors of pain-related disability. We document significantly greater peak gray matter density in individuals with LBP in the primary somatosensory cortex, angular gyrus, and the midcingulate cortex. Pain-related disability positively correlated with average gray matter density in the posterior cingulate cortex. The most robust predictors of disability were average gray matter in the posterior cingulate, pain intensity, and pain-related fear. We demonstrate that in young adults, persistent LBP, and pain-related disability, are linked with structural neuroplasticity in regions forming part of the brain network termed the pain matrix. In contrast with studies of LBP in older adults, our findings of increased rather than decreased gray matter in young adults with LBP suggest that gray matter may increase initially in response to nociceptive pain.

## INTRODUCTION

Structural brain plasticity is a dynamic process of adaptation that is shaped by an individual’s experience and environment.[27] Structural reorganization in the brain in response to experience in turn influences neural function and future behavior.[27] These principles of neuroplasticity are hypothesized to be mechanisms underlying behavioral, emotional, cognitive, and sensory features of many clinical disorders, including persistent pain.[31]

Structural neuroplasticity in gray matter following an initial episode of pain may contribute to the persistence of low back pain (LBP).[60] Multiple studies have identified differences in gray matter in individuals with LBP compared with pain-free individuals. Initial evidence suggested that persistent LBP primarily associates with decreased gray matter in cortical and sub-cortical brain regions involved in pain perception.[3,45] More recent reviews indicate region-specific findings of both increased and decreased gray matter in individuals with persistent LBP.[24,29,60,62] It has been reported that adaptations in gray matter evolve over time and that they associate with duration of pain.[3,30] However, some studies showing time-dependent alterations in gray matter morphology have been limited by small sample sizes, and their findings may be specific to older participants with multi-decade histories of pain.[3,6,45]

Most individuals with persistent LBP experienced their first episode of pain as a young adult or adolescent.[25] It is critical therefore to determine the neuroplasticity associated with persistent pain specifically in young adults. It is also important to establish if development of neuroplasticity in individuals with LBP is time- or age-dependent. This knowledge may facilitate early identification of individuals who have neural signatures that place them at risk of a decades-long relationship with LBP. To our knowledge, brain morphology has not been evaluated in young adults who are still early in the time-course of LBP. However, studies with participants in their thirties suggest that plasticity in gray matter in cortical sensorimotor regions such as the primary somatosensory cortex may have a key role in pain persistence early in adulthood.[28,55]

The influence of LBP on an individual’s life may be best assessed by the extent of their pain-related disability. Pain-related disability results from the interaction of the behavioral, emotional, cognitive, and sensory aspects of pain.[64] It is not clear if there are brain regions or networks that directly influence the extent of pain-related disability that an individual with LBP experiences. The severity of back pain symptoms is known to be one of the strongest predictors of current and future disability in individuals with LBP.[20] Pain-related fear is also an important contributor to the degree of disability associated with LBP.[20,64] Therefore structural neuroplasticity may also influence disability via associations with clinical pain characteristics such as symptom intensity or pain-related fear.

This study had two aims. The first was to determine if gray matter in cortical sensorimotor regions differs in young adults with a history of persistent low back pain compared to young adults with no history of back pain. The second aim of the study was to identify if sensorimotor gray matter is an independent predictor of pain-related disability, or associates with clinical characteristics known to be contributors to disability, in young adults with a history of LBP.

## METHODS

Eighty-five young adults between the ages of 18 and 35 years (mean age 22.6 ± 3.5 years) were recruited to the study. Fifty-five individuals had a history of low back pain, and 30 controls had no history of back pain. Participants were recruited via flyers posted at multiple college campuses and in gyms/athletic facilities, as well as via social media and by word of mouth. All volunteers provided written informed consent and the study was approved by the Institutional Review Board of Chapman University.

For inclusion in the group with LBP, participants had to have at least a one-year history of pain that was primarily localized between the lower rib and the gluteal fold. Pain could be unilateral, bilateral, or central. Individuals who reported proximal leg pain in addition to lower back pain were included if there were no signs or symptoms of a radiculopathy. Individuals in the group with LBP had to have experienced at least one episode of pain in the preceding six months. An episode was defined as a period when the symptoms were severe enough to limit functional activity and that lasted for a minimum of 24 hours.[51] Participants with LBP had to be asymptomatic at the time of data collection. This was defined as a current pain level of less than 5/100 on a visual analogue scale.[49] For inclusion in the control group, individuals had to have no history of back pain that required medical care or more than three days of activity limitation, and no other active pain condition. Participants were excluded from both groups if they had any of the following characteristics: history of other chronic or widespread pain conditions; history of significant musculoskeletal or neurological disorders; history of systemic inflammatory disease; history of brain injury or spinal surgery; history of significant traumatic, inflammatory, or structural spinal pathology including high-grade spondylolisthesis or radiculopathy; scoliosis greater than 30 degrees; history of smoking; being involved in current litigation related to LBP; and contraindications to MR imaging. All participants reported right limb preference, quantified using the Lateral Preference Inventory,[11] except for two participants who were ambilateral.

### Physical activity level and physical quality of life

In order to assess the potential for activity level to influence gray matter morphology and pain-related disability, typical activity level was quantified with the Physical Activity Scale (PAS). [1] This is a self-report measure that quantifies physical activity over a typical 24-hour period. Participants identify the amount of time they spend in activities that vary from sedentary to highly physically demanding. A total score is calculated by multiplying the metabolic equivalents (METs) for each type of activity by the duration of the activity (MET-time). The Physical Activity Scale has been validated in comparison with exercise diaries and with cardiovascular fitness.[1,2] Quality of life in the physical health domain was asessed using the Brief Version of the World Health Organization Quality of Life assessment physical health subscale (WHOQOL-PH).

### Clinical characteristics in individuals with LBP

In individuals with LBP pain-related disability was evaluated using the modified Oswestry Disability Index (ODI); a valid and reliable self-report measure. Extent of disability is quantified as a percentage.[13] Three clinical characteristics with the potential to contribute to pain-related disability were also assessed: pain duration, pain intensity, and pain-related fear. Pain duration was quantified as the time since first onset of symptoms. Pain intensity was defined as the severity of pain during a typical episode, and was quantified using a 0 to 100mm visual analogue scale, with the ends points of the line representating “no pain” and “worst pain imaginable”. Pain-related fear was quantified with the Fear Avoidance Beliefs Questionnaire (FABQ).[59] The fear avoidance model encompasses pain-related distress and anxiety, and the cognitive appraisal of the risk of pain and potential harm of physical or work-related activities.[17,20] The FABQ is a valid and reliable measure of the fear avoidance construct.[19] Both the work and the physical activity subscales of the FABQ are predictive of clinical outcomes in individuals with LBP.[18] In this research we focused on the physical activity subscale of the FABQ (FABQ-PA) as most of the young adults in our study were not in full time employment, and because of previous evidence of relationships between pain, disability, and the physical activity subscale of the FABQ.[19,22,35,50,64]

### Image acquisition and preprocessing

Gray matter was quantified using voxel-based morphometry.[5,65] T1-weighted structural images were acquired with a 3-T Siemens Prisma scanner (Siemens Medical Solutions USA, Inc, Malvern, PA) using a 32-channel head coil. The 3D-MPRAGE sequence had the following parameters: TR 2400 ms, TE 4.16 ms, and isotropic voxel size = 0.7mm^3^. Image preprocessing was conducted using the Computational Anatomy Toolbox (CAT12 version 12.7,[16]) implemented in SPM12 (version 7771) in MATLAB (version 2018b, MathWorks, Natick, MA). Images were bias corrected, affine-registered, and then segmented using the SPM Unified Segmental Approach.[4,5] They were registered to the CAT default Geodesic Shooting template using Shooting Registration.[16] The normalized and modulated images were then smoothed in SPM12 using a full width at half maximum 8mm Gaussian kernel. This resulted in images with gray matter density quantified in arbitrary units.

Cortical sensorimotor regions of interest (ROIs) were identified *a priori*. We selected cortical regions involved in sensorimotor function that were associated with back pain in previous studies.[21,32,48] We investigated the following ROIs: pre- and post-central gyri, supplementary motor area, cingulate cortex (subdivided into anterior cingulate, midcingulate, and posterior cingulate cortices), superior parietal lobule, inferior parietal lobule, and supramarginal gyrus/parietal operculum. Region of interest masks were constructed using the Automated Anatomical Labelling Atlas (AAL,[43,54]) in WFU Pickatlas (Version 3.05, https://www.fmri.wfubmc.edu/downloads,[33,34]). The average gray matter in each region of interest for each individual was extracted using the Neuromorphometrics atlas in CAT12 and was exported for additional analysis.

### Statistical analyses

Sample size was determined *a priori* by a power analysis using published literature that indicated that a sample size of 26 per group would provide >80% power to detect a difference in S1/M1 gray matter density between individuals with and without LBP with an effect size of at least 0.8.[45]

#### Group comparisons

We tested for localized group differences in gray matter within ROIs using voxel-wise analyses, and tested for more distributed group differences in gray matter using the average gray matter within each ROI. Regional group differences in peak gray matter density in each ROI were determined using the voxel-wise general linear model in SPM12. Gray matter volume was normalized to total intracranial volume (TIV) using global scaling. A threshold mask with an absolute value of 0.1 was applied to minimize edge effects at boundaries between tissue types. Age and sex were included as covariates. We tested for group differences using a family-wise error rate corrected threshold of p < 0.05 and minimum voxel extent of 5 voxels. We examined the contrasts LBP > control and control > LBP in each ROI.

To compare average gray matter in the ROIs we used ANCOVA with age, sex, and TIV as covariates (SPSS version 28.0.1.1, IBM®, Armonk, USA).

#### Predictors of pain-related disability in individuals with LBP

Gray matter density predictors of pain-related disability were identified using Pearson correlation coefficients. We adjusted for age, sex, and TIV in all analyses. The primary analysis first identified ROIs that uniquely explained variance in pain-related disability by determining associations between average gray matter density and pain-related disability, after adjustment for levels of pain intensity, pain duration, and pain-related fear. Then, the secondary analysis identified ROIs that contributed to disability via an association with the other relevant clinical characteristics by determining associations between average gray matter density and pain intensity, pain duration, and pain-related fear. Data were checked to confirm that they met the assumptions for bivariate normality and were log-transformed as needed. As these analyses were exploratory and were used to identify variables to include in the multivariate model, we did not correct for multiple comparisons.

We then examined multivariate predictors of pain-related disability using the ROIs that were significantly correlated with pain-related disability, or that were correlated with the other clinical characteristics. We assessed linear regression models, including age, sex, and TIV as covariates in all models. The regression models were checked for normal distribution of residuals, multivariate outliers, and collinearity of independent variables.

## RESULTS

Participant demographics and clinical characteristics of the participants in the group with a history of LBP are shown in Table 1. Imaging was not completed in three participants due to scheduling challenges (two individuals with LBP) and anxiety in the scanner (one control participant). Participants with a history of LBP were slightly younger than those in the control group (21.9 ± 3.1 years and 23.7 ± 4.0 years respectively). There was no difference in the proportion of males and females in the groups. Participants maintained moderate levels of physical activity[2,46] and this did not differ between groups. Individuals with a history of LBP reported significantly poorer physical quality of life than those without LBP (WHOQOL-PH 26.9 ± 4.3 and 29.7 ± 3.8 respectively).

**Table 1.**
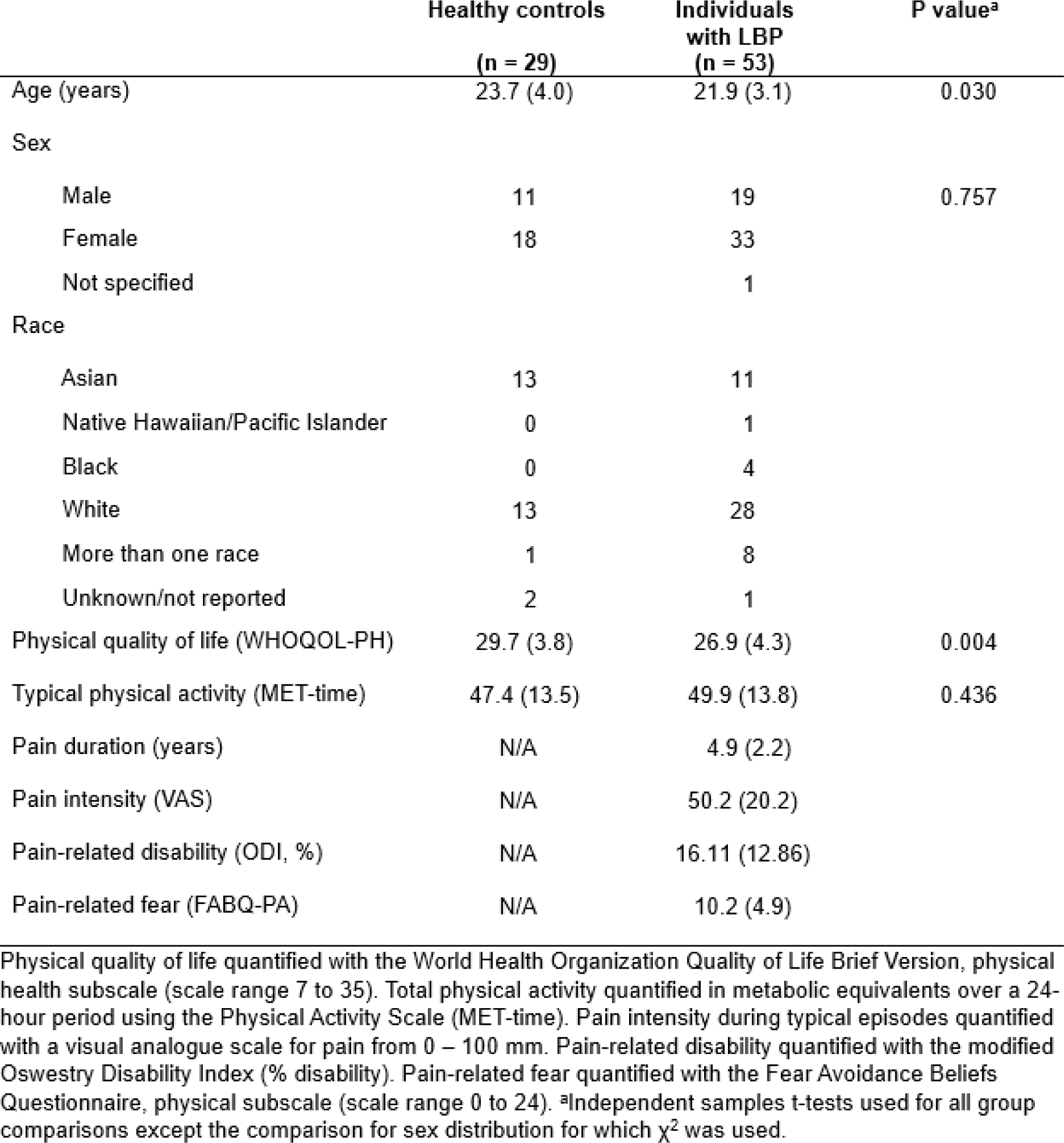
Group demographics of both groups and clinical characteristics of individuals with a history of low back pain.

Pain-related disability was heterogeneous in the group with LBP. The average score was 16.11%, indicating minimal disability. Individual scores ranged from 0% (no disability) to 56% (severe disability). [13] Average duration of symptoms in individuals with LBP was 4.9 ± 2.2 years. Sixty-five percent of the participants reported a recurrent/episodic pattern of pain symptoms while the remainder reported experiencing symptoms on at least half of the days in the prior six-month period, consistent with a commonly-used criterion for chronic pain.[12] Participants reported a moderate average symptom intensity during episodes of 50.2 ± 20.2/100.[8] Average pain-related fear (FABQ-PA, physical activity domain) was 10.2 (± 4.9). This is below the threshold of 15 that is considered to be predictive of a poor clinical outcome.[18]

### Group comparisons

Localized group differences in peak gray matter density in the selected ROIs are shown in Figure 1 and Table 2. All statistically significant group differences were evident in the right hemisphere. Individuals with a history of LBP demonstrated greater gray matter density than the control group in the post-central gyrus (approximating primary somatosensory cortex), inferior parietal lobule (specifically the angular gyrus), and the right midcingulate cortex. There were no areas in which the control group had greater gray matter density than the group with LBP.

**Figure 1.**
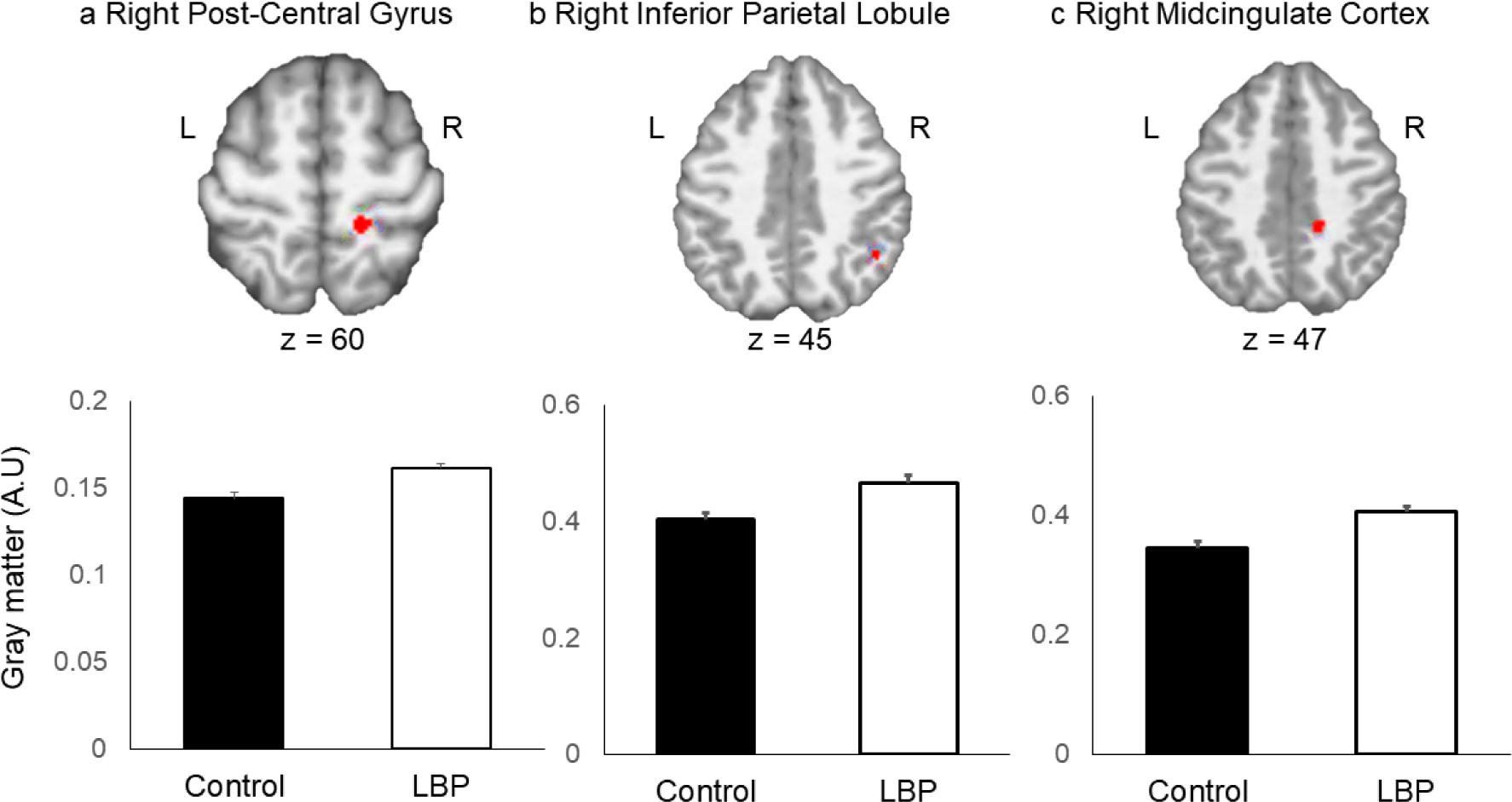
Differences in peak gray matter between the individuals with a history of LBP and the back-healthy control group. There was significantly greater gray matter in the group with LBP in the following ROIs: a) Right Post-Central Gyrus; b) Right Inferior Parietal Lobule; and c) Right Midcingulate Cortex. For the purposes of visualization, the clusters shown (top) were extracted with minimum extent of 5 voxels and height threshold of 0.001. Values shown (below) are gray matter density at the peak voxel in the ROI, significant at the FWE-corrected p < 0.05. A.U = arbitrary units. Error bars are standard errors.

**Table 2.** Group differences in peak gray matter density in sensorimotor regions of interest.

| Region of interest | BA | Z-score | P<sub>FWE</sub> - corrected | MNI coordinates |  |  |
| --- | --- | --- | --- | --- | --- | --- |
|  |  |  |  | x | y | z |
| Contrast: LBP > Control |  |  |  |  |  |  |
| R Post-Central Gyrus | 3B | 4.18 | 0.009 | 17 | -35 | 60 |
| R Inferior Parietal Lobule | 39 | 3.50 | 0.029 | 45 | -51 | 45 |
| R Midcingulate Cortex | 31 | 4.13 | 0.011 | 17 | -39 | 47 |
| Contrast: Control > LBP |  |  |  |  |  |  |
| No suprathreshold voxels |  |  |  |  |  |  |

*Threshold $p < 0.05$ FWE corrected at the peak level. Minimum cluster size of 5 voxels, reported in MNI space. Z-scores of peak voxels are shown. Age and sex were included as covariates and gray matter was normalized to total intracranial volume. Locations of significant local maxima in the ROI are listed and labeled using the Automated Anatomical Labelling Atlas (AAL). Where applicable cytoarchitectonic areas (Brodmann areas, BA) are also provided and defined using the SPM Anatomy Toolbox and the Biomed MNI2Tal tool.*

The average gray matter density in each ROI, adjusted for age, sex, and TIV, did not differ between groups (p > 0.05 for all comparisons, see Supplemental Information).

### Bivariate associations between average gray matter density and pain-related disability in individuals with LBP

Significant bivariate associations between average gray matter density and pain-related disability are shown in Figure 2 and Table 4. Data for all correlation analyses are shown in the Supplemental Information.

**Figure 2.**
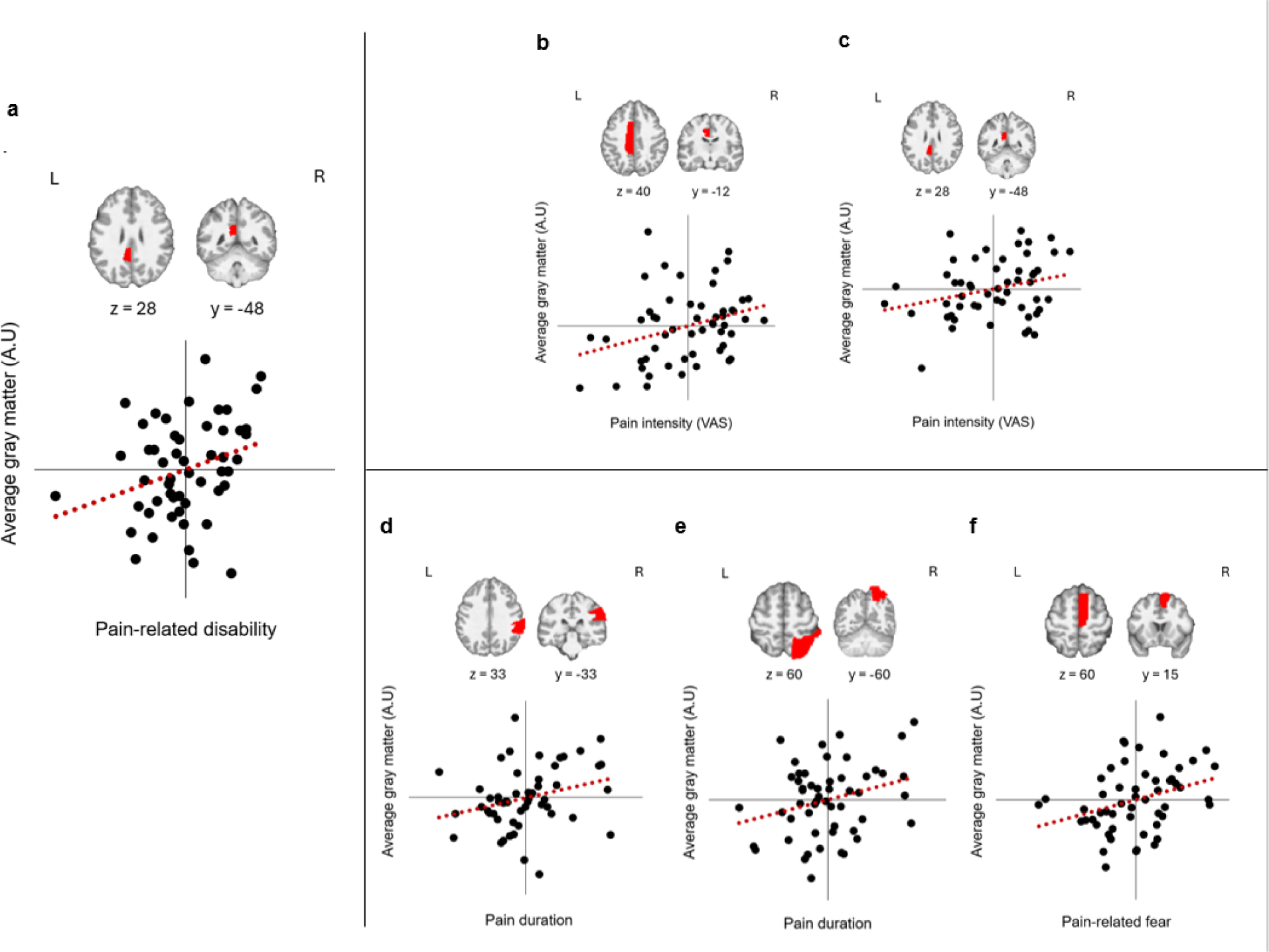
Associations between pain-related disability and average gray matter density in sensorimotor regions of interest (a), and between the other clinical characteristics and average gray matter density (b – f). Note that the plots show partial correlations using residuals obtained after adjusting for covariates: a) pain-related disability (ODI) and left posterior cingulate cortex; b) pain intensity and left midcingulate cortex; c) pain intensity and left posterior cingulate cortex; d) pain duration and right supramarginal gyrus; e) pain duration and right superior parietal lobule; f) pain-related fear (FABQ-PA) and right supplementary motor area.

Pain-related disability was positively correlated with average gray matter density in the left posterior cingulate cortex (r = 0.292, p = 0.046). The secondary analyses of bivariate associations between average gray matter density and clinical characteristics demonstrated that pain intensity was significantly positively correlated with average gray matter density in the left midcingulate cortex (r = 0.316, p = 0.025) and left posterior cingulate cortex (r = 0.280, p = 0.043). Duration of symptoms was positively correlated with average gray matter density in the right supramarginal gyrus (r = 0.281, p = 0.048) and the right superior parietal lobule (r = 0.285, p = 0.045). Pain-related fear was positively correlated with average gray matter density in the right supplementary motor area (r = 0.339, p = 0.019).

### Multivariate predictors of pain-related disability

Multivariate predictors of pain-related disability in individuals with LBP were explored using two models (Table 4). Model 1 examined the clinical determinants of pain-related disability (ODI score). Age and sex were included in the model. Greater pain intensity and higher pain-related fear (FABQ-PA scores) were both statistically significant predictors of higher pain-related disability (adjusted R^2^ 0.215, F (5, 47) = 3.853, p = 0.005).

Model 2 examined average gray matter in ROIs and the clinical determinants of pain-related disability (ODI score). Age, sex, and TIV were included in the model. ROIs were included in the model if they had a bivariate association with pain-related disability, or with one of the clinical characteristics (Table 3). Due to multicollinearity between average gray matter in the left midcingulate cortex and the left posterior cingulate cortex, we examined two versions of Model 2, with either the left midcingulate or the left posterior cingulate included in addition to the other variables. The model with the left posterior cingulate explained the greatest amount of variance. In addition to pain intensity and FABQ-PA scores, greater pain-related disability was significantly predicted by higher average gray matter density in the left posterior cingulate cortex (adjusted R^2^ 0.241, F (10, 42) = 2.655, p = 0.013). For the final versions of both models, the variance inflation factor for all variables was below 4, indicating acceptable collinearity between the independent variables.[37]

**Table 3.**
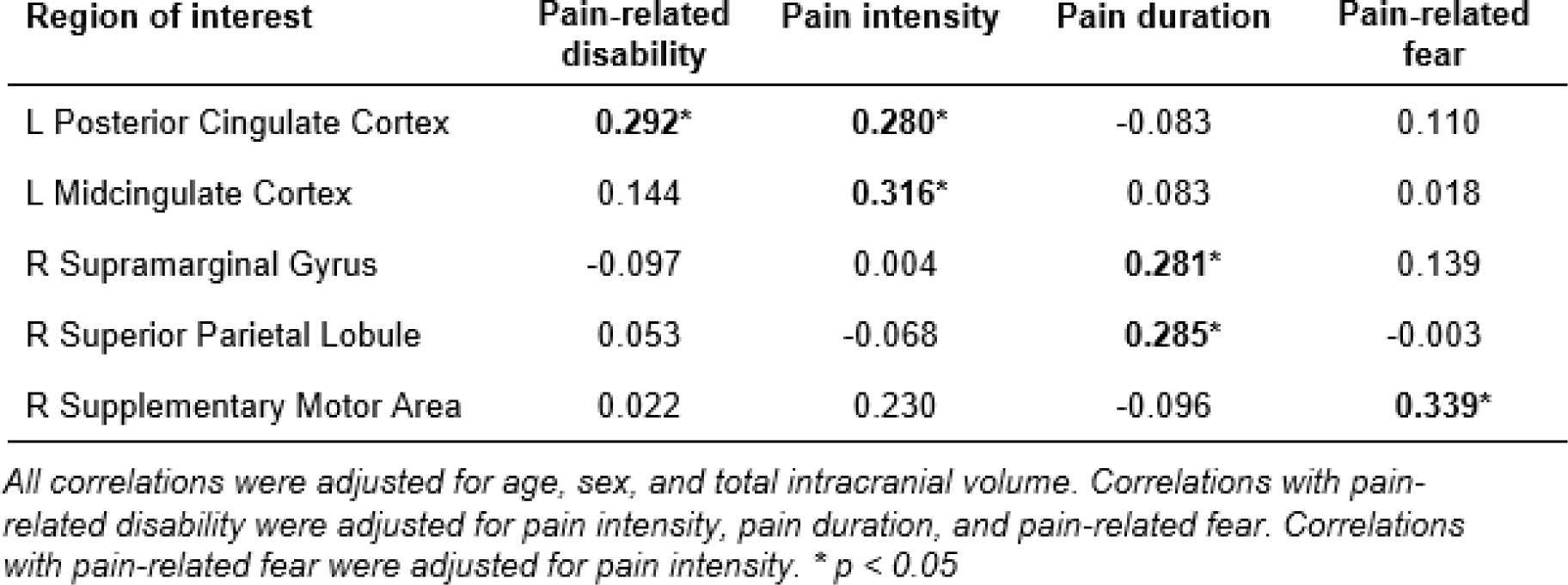
Bivariate associations between average gray matter density in sensorimotor regions of interest, pain-related disability, and the other clinical characteristics.

**Table 4.** Linear regression models. Model 1 tested associations between clinical characteristics and pain-related disability. Model 2 tested associations between average gray matter density, clinical characteristics, and pain-related disability. Unstandardized coefficients (B), standard errors (SE B), and standardized coefficients (**β)** are shown. Significant predictors are indicated with standardized coefficients in bold. The model that explained the greatest variance in pain-related disability included pain intensity, pain-related fear, and average gray matter in the left posterior cingulate cortex.

|  | Model 1 |  |  | Model 2 |  |  |
| --- | --- | --- | --- | --- | --- | --- |
| | B | SE B | $\beta$ | B | SE B | $\beta$ |
| Pain intensity | 0.006 | 0.002 | **0.412\*\*** | 0.005 | 0.002 | **0.339\*** |
| Pain duration | 0.025 | 0.020 | 0.173 | 0.033 | 0.022 | 0.230 |
| Pain-related fear | 0.021 | 0.009 | **0.322\*** | 0.021 | 0.010 | **0.325\*** |
| Avg L Post Cingulate Cortex | - | - | - | 0.291 | 0.123 | **0.534\*** |
| Avg R Sup Parietal Lobule | - | - | - | -0.009 | 0.062 | -0.026 |
| Avg R Supramarginal Gyrus | - | - | - | -0.055 | 0.053 | -0.143 |
| Avg R Suppl Motor Area | - | - | - | -0.095 | 0.123 | -0.193 |

*Model 1 included age and sex. Model 2 included age, sex, and total intracranial volume. \* $p < 0.05$ . \*\* $p < 0.01$*

Pain intensity and average gray matter density in the left posterior cingulate were significantly correlated with each other and were both predictors of disability. We conducted a post-hoc mediation analysis to determine if increased average gray matter in the posterior cingulate is a mediator of the relationship between pain intensity and disability. Further details of this analysis are provided in the Supplemental Information. We found that the strength of the relationship between pain intensity and disability was not significantly reduced after accounting for effects of gray matter density (indirect effect β=0.080 (95% CI -0.007 to 0.199), Sobel’s test t = 1.473, p = 0.141). Therefore, a mediation role for gray matter density in the posterior cingulate in the relationship between pain intensity and pain-related disability was not supported. following ROIs: a) Right Post-Central Gyrus; b) Right Inferior Parietal Lobule; and c) Right Midcingulate Cortex. For the purposes of visualization, the clusters shown (top) were extracted with minimum extent of 5 voxels and height threshold of 0.001. Values shown (below) are gray matter density at the peak voxel in the ROI, significant at the FWE-corrected p < 0.05. A.U = arbitrary units. Error bars are standard errors.

## DISCUSSION

Our study demonstrates that differences in gray matter in cortical sensorimotor areas are evident early in the time-course of persistent LBP in young adults. We also show for the first time that gray matter density is an independent predictor of the pain-related disability experienced by young adults with a history of back pain.

The areas of increased gray matter in individuals with LBP in our study were in brain regions subserving functions relevant to sensory processing. The location of increased gray matter in S1 was consistent with the medial somatotopic representation of the trunk.[26] Kim et al.,[26] found increased gray matter in the same location in their study of individuals with persistent LBP. Their S1 ROI was determined experimentally using tactile sensation in the lower back, and they reported that greater gray matter in this region was associated with more impaired two-point discrimination in the lower back. The cluster of voxels with increased gray matter that we found in the cingulate cortex was located at the border of the posterior midcingulate cortex and the dorsal posterior cingulate cortex.[57,58] This part of the cingulate cortex receives substantial afferent input from the parietal cortex and utilizes this multisensory information to orient the body in space, to guide the motor response to nociception and other noxious stimuli, and for spatial navigation.[41,57] Both the somatosensory and cingulate cortices form part of the network of brain regions that have been termed the “pain matrix”.[40,61] The primary somatosensory cortex is part of the primary cortical pain matrix, with a role in the localization and perception of pain. The midcingulate and posterior cingulate cortices are part of the tertiary cortical pain matrix which cognitively appraises sensory information and initiates a behavioral response to it.[61]

The increased gray matter in the caudal inferior parietal lobule in our participants was in the angular gyrus (Brodmann 39). This region is associated with processing of multimodal sensory input, orientation of attention to sensory input, and motor preparation.[10,42] In the right hemisphere, damage to the angular gyrus results in altered body image and impaired conscious recognition of the body in space (sometimes termed body schema).[7,56] Our findings are consistent with behavioral evidence that suggests that individuals with persistent LBP, and those with other chronic pain disorders, have altered body schema.[9,56] The right angular gyrus is also part of the central executive network, with structural and functional connections to the right dorsolateral prefrontal cortex. This indicates a role in decision making and task control. [47] Alterations in the structure and function of the central executive network in individuals with LBP may therefore provide a link between the perception and sensory processing of pain and the functional limitation/disability associated with it.[40]

Our results in young adults with a history of LBP add clarity to findings from previous studies. Importantly, the average age of our participants was almost 20 years younger than that of individuals with mechanical LBP included in previous studies of gray matter morphology,[38,62], and the average duration of symptoms in our cohort was half that of the majority of existing studies.[63] A previous study with participants close in age to our own reported multiple regions - including S1, S2, and M1 - where increased gray matter distinguished individuals with persistent LBP from a back-healthy control group.[55] Yang et al.’s recent meta-analysis [62] found that gray matter in the left post-central gyrus was consistently increased across studies in individuals with persistent LBP. In contrast, Henn et al., [24] synthesized findings from studies of several chronic pain conditions, predominantly in middle-aged participants, and did not identify any regions where individuals with pain had greater gray matter. The difference in the results of these two meta-analyses may reflect heterogeneity in the neuroplasticity associated with different chronic pain conditions, and disorder-specific somatotopy in brain morphology.[24] Areas of decreased gray matter in individuals with back pain are most commonly reported to include the anterior cingulate and midcingulate cortices, primary and secondary somatosensory cortices, the insula, thalamus, superior frontal gyrus, striatum and middle occipital gyrus.[24,36,62,63]

Based on our findings and those of earlier studies we speculate that in the early stages of persistent LBP, gray matter increases due to an initial neuroplastic response to greater usage and activation of areas associated with nociception and pain processing. Mechanisms underlying increased gray matter may include increased size/density of neurons, dendrites, synapses, and neurotransmitter receptors, increased local brain metabolism, increased extracellular water content, and vascular volume.[39,44,52] Over prolonged periods of time, gray matter may then gradually atrophy due to excitotoxity and neurodegeneration as a result of overuse.[3,30] In addition, over time the locus of pain processing may shift away from the primary pain matrix to regions such as the anterior cingulate that are more associated with the attentional and affective aspects of the pain experience.[23] We cannot exclude an alternative hypothesis that increased gray matter morphology in sensorimotor regions may be a predisposition to and not an effect of an initial episode of LBP.[45] However, our hypothesis that gray matter increases initially in response to nociceptive pain is supported by a study that demonstrated that inducing experimental nociceptive pain over the course of eight days in healthy participants resulted in increased gray matter in regions including the midcingulate cortex, somatosensory cortex and the inferior parietal lobule.[53] This hypothesis is also supported by our findings of a positive correlation between average gray matter density in the supramarginal gyrus and superior parietal lobule and pain duration.

Our study aimed to determine whether gray matter neuroplasticity contributes to pain-related disability, either directly, or via association with pain intensity, pain duration, or pain-related fear. The model that explained the greatest amount of variance in disability in our participants included the participant’s typical pain intensity, the extent of their pain-related fear, and average gray matter in the left posterior cingulate cortex. The contribution of cingulate gray matter to pain-related disability was independent of pain intensity or pain-related fear. The posterior cingulate cortex is known to subserve a variety of complex functions.[15] Sub-regions of the posterior cingulate connect with brain areas associated with somatosensation, memory, executive control, and the limbic system.[15,58] In particular, the dorsal portion of the posterior cingulate associates with executive control networks, while the ventral portion is part of the default mode network.[15] Therefore, the contribution of the posterior cingulate to the disability associated with persistent pain may include assessment of the personal relevance of sensory information, cognitive appraisal of the pain experience, and behavioral decision making. [14,38,58] The fact that the posterior cingulate was the only region to independently associate with pain-related disability in our study suggests that pain appraisal may be more important than nociception or emotion in shaping behaviors that lead to pain-related disability.[38]

In addition to the posterior cingulate cortex, our exploratory correlation analyses identified several regions where average gray matter density was significantly associated with clinical characteristics. Greater typical pain intensity associated with greater average gray matter in the left midcingulate cortex. The anterior midcingulate is activated by nociceptive pain. [38] It is associated with negative pain affect, anticipation of pain, and pain sensitivity as part of the tertiary pain matrix[61], and with selection of motor responses in the context of reward and outcomes associated with previous motor behaviors.[41,52,57]. Higher pain-related fear associated positively with greater average gray matter in the right supplementary motor area. This finding may support a role for the SMA for developing a behavioral (motor) response to perceived pain.[14] In contrast to existing studies demonstrating worsening gray matter atrophy over time[3,6] the only relationship we observed between gray matter and duration of symptoms was the positive association between longer duration of symptoms and greater average gray matter in the secondary association cortex (superior parietal lobule) and the supramarginal gyrus.

We acknowledge some limitations to this research. Our study was cross-sectional and involved participants who already had at least a one-year history of pain. Therefore, additional studies will be needed to determine if young adults with greater gray matter neuroplasticity have more adverse symptom trajectories, and to examine how characteristics of gray matter neuroplasticity evolve across the time-course of back pain. Further research clarifying the relationship between posterior cingulate neuroplasticity and pain-related disability will be needed to identify a neurological model or phenotype for disability development that can then be tested empirically. Additionally, we did not quantify potential socioeconomic contributors to pain-related disability such as income or social support.

### Conclusion

Our findings suggest that gray matter neuroplasticity is evident even in young, minimally disabled adults with a relatively benign pattern of back pain symptoms. Neuroplasticity in the posterior cingulate cortex may be an important determinant of the disability associated with back pain. Gray matter density may therefore have utility as a biomarker of neural changes related to development of LBP. This finding also offers the potential for novel early interventions targeting gray matter neuroplasticity in young adults with persistent pain.

## Data Availability

All data produced in the present study are available upon reasonable request to the authors

## Notes

### Competing Interest Statement

The authors have declared no competing interest.

### Funding Statement

This study was funded by the Eunice Kennedy Shriver National Institute of Child Health and Human Development. Award # K01HD092612.

### Author Declarations

The Institutional Review Board of Chapman University gave ethical approval for this work.

