## Supplemental information for "Gray matter morphology and pain-related disability in young adults with low back pain"

|  | **Healthy controls** | **Individuals with LBP** | **P value** |
| --- | --- | --- | --- |
| R Medial Pre-Central Gyrus | 2.46 (0.29) | 2.60 (0.37) | 0.060 |
| L Medial Pre-Central Gyrus | 2.38 (0.41) | 0.25 (0.31) | 0.290 |
| R Medial Post-Central Gyrus | 1.15 (0.20) | 1.20 (0.18) | 0.087 |
| L Medial Post-Central Gyrus | 1.21 (0.74) | 1.09 (0.18) | 0.277 |
| R Supplementary Motor Area | 5.91 (0.63) | 6.00 (0.65) | 0.977 |
| L Supplementary Motor Area | 6.50 (1.93) | 6.32 (0.75) | 0.483 |
| R Anterior Cingulate Cortex | 4.08 (0.51) | 4.00 (0.64) | 0.251 |
| L Anterior Cingulate Cortex | 5.50 (1.14) | 5.61 (0.76) | 0.820 |
| R Midcingulate Cortex | 4.57 (0.47) | 4.64 (0.52) | 0.474 |
| L Midcingulate Cortex | 4.55 (0.70) | 4.67 (0.44) | 0.456 |
| R Posterior Cingulate Cortex | 4.07 (0.53) | 3.98 (0.48) | 0.149 |
| L Posterior Cingulate Cortex | 4.26 (0.75) | 4.36 (0.58) | 0.585 |
| R Superior Parietal Lobule | 9.98 (0.94) | 10.24 (0.95) | 0.442 |
| L Superior Parietal Lobule | 10.08 (1.62) | 10.41 (1.03) | 0.319 |
| R Angular Gyrus | 10.09 (1.11) | 10.46 (1.12) | 0.350 |
| L Angular Gyrus | 8.80 (1.55) | 9.23 (0.94) | 0.114 |
| R Supramarginal Gyrus | 6.31 (0.73) | 6.61 (0.83) | 0.154 |
| L Supramarginal Gyrus | 6.79 (0.03) | 6.94 (0.94) | 0.221 |

**Table S1**. Group comparisons of average gray matter density (in cm^3^) in cortical sensorimotor regions of interest. Age, sex, and total intracranial volume were included as covariates. Group mean (± standard deviation).

**Table S2.** Bivariate associations between average gray matter density in sensorimotor regions of interest, pain-related disability, and the other clinical characteristics.

| **Region of interest** | **Pain-related disability** | | **Pain intensity** | **Pain duration** | **Pain-related fear** |
| --- | --- | --- | --- | --- | --- |
| R Medial Pre-Central Gyrus | | 0.251 | 0.147 | -0.030 | 0.019 |
| L Medial Pre-Central Gyrus | | 0.218 | 0.037 | -0.054 | 0.052 |
| R Medial Post-Central Gyrus | | 0.139 | 0.051 | -0.107 | 0.107 |
| L Medial Post-Central Gyrus | | 0.035 | 0.230 | 0.041 | -0.093 |
| R Supplementary Motor Area | | 0.022 | 0.230 | -0.096 | **0.339*** |
| L Supplementary Motor Area | | 0.204 | 0.180 | -0.053 | 0.190 |
| R Anterior Cingulate Cortex | | 0.165 | 0.081 | -0.015 | -0.091 |
| L Anterior Cingulate Cortex | | 0.148 | 0.107 | 0.141 | 0.021 |
| R Midcingulate Cortex | | 0.030 | 0.177 | 0.008 | -0.005 |
| L Midcingulate Cortex | | 0.144 | 0.316* | 0.083 | 0.018 |
| R Posterior Cingulate Cortex | | 0.240 | 0.154 | -0.177 | 0.181 |
| L Posterior Cingulate Cortex | | **0.292*** | **0.280*** | -0.083 | 0.110 |
| R Superior Parietal Lobule | | 0.053 | -0.068 | **0.285*** | -0.003 |
| L Superior Parietal Lobule | | -0.154 | 0.169 | -0.115 | -0.084 |
| R Angular Gyrus | | -0.153 | 0.095 | 0.070 | 0.060 |
| L Angular Gyrus | | -0.180 | -0.036 | 0.120 | 0.094 |
| R Supramarginal Gyrus | | -0.097 | 0.004 | **0.281*** | 0.139 |
| L Supramarginal Gyrus | | -0.067 | 0.019 | 0.162 | 0.034 |

**Mediation analysis**

A post-hoc mediation analysis was conducted to determine if posterior cingulate gray matter (mediator, M) mediated the relationship between pain intensity (independent variable, X) and disability (dependent variable, Y), using the PROCESS macro in SPSS (simple mediation, model 4).[24] Direct and indirect effects of the independent variables on ODI score were tested using bootstrapping (5000 samples) to calculate 95% confidence intervals of the estimates. The significance of the indirect effect was further tested using the Sobel test.[39] Age, sex, and TIV were included as covariates in the mediation model. We found that the strength of the relationship between pain intensity and disability was not significantly reduced after accounting for effects of gray matter density (indirect effect β=0.080 (95% CI -0.007 to 0.199), Sobel’s test t = 1.473, p = 0.141, Figure S1). Therefore, a mediation role for gray matter density in the posterior cingulate in the relationship between pain intensity and pain-related disability was not supported.

**Figure S1**. The effect of pain intensity on pain-related disability (ODI score) was not significantly reduced when the indirect effect of posterior cingulate gray matter was removed. Thus, the model does not support mediation. Standardized coefficients are shown for each pathway. * indicates p<0.05.

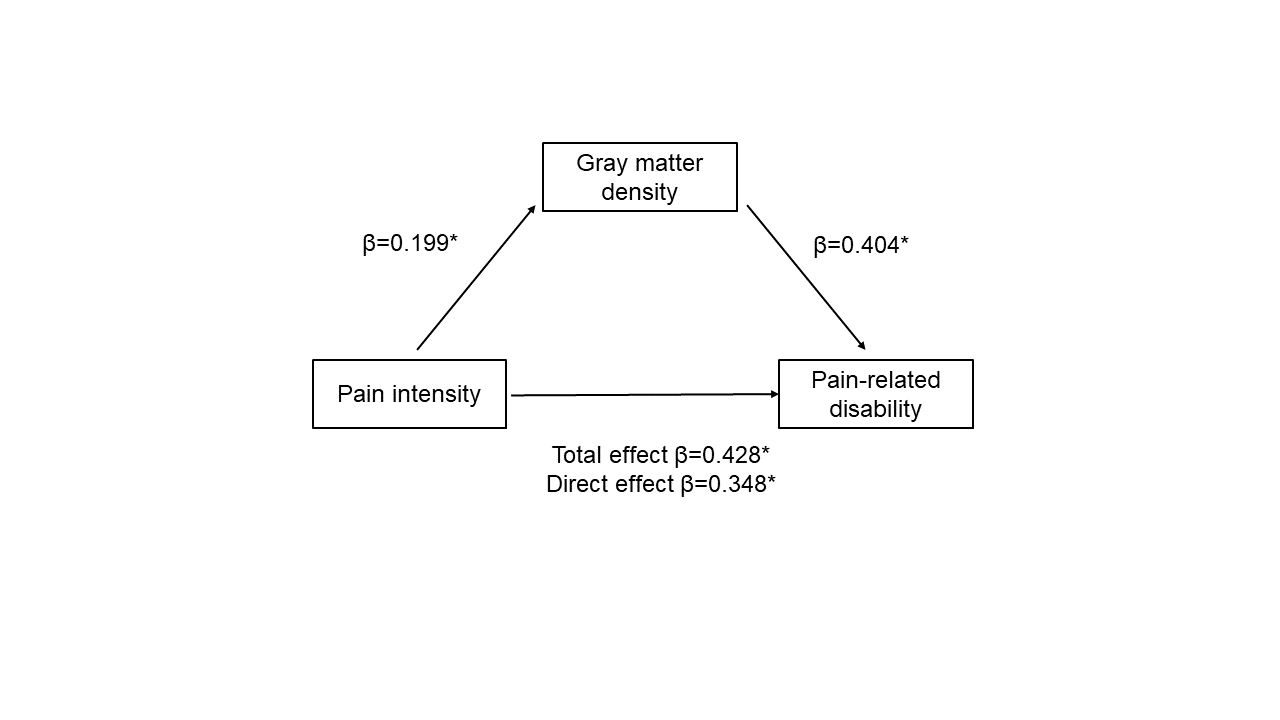
